# Evaluating the Therapeutic Effects of Amino Acid Treatment on Vaso-Occlusive Pain in Sickle Cell Disease: A Systematic Review and Meta-Analysis Protocol

**DOI:** 10.1101/2024.08.08.24311691

**Authors:** Bohan Zhang, Ciaran Bubb, Sophie Yao, Vivian Dong, Priyal Patel, Aanya Shahani, Katie Lobner, Oluwakemi Badaki-Makun

## Abstract

2

**Introduction:** Sickle Cell Disease (SCD) affects over 100,000 individuals in the United States and 20 million globally, causing acute and chronic pain. The disease is characterized by misshapen red blood cells caused by mutations in beta-hemoglobin genes. SCD leads to multiorgan damage, chronic anemia, and severe pain crises, with a median life expectancy of 43 years. Current treatments involve opioids, blood transfusions, and hydroxyurea. Amino acids, especially L-Glutamine, have shown promise in managing SCD pain. This systematic review aims to comprehensively analyze the effects of amino acid treatments on vaso-occlusive pain crises in SCD patients.

**Methods:** Following the Cochrane Handbook and PRISMA-P guidelines, this systematic review will include studies comparing amino acid treatment to placebo or standard care for SCD pain crises. Eligible studies of all age ranges, settings, and amino acid types will be considered. A comprehensive search strategy will be employed in PubMed, Embase, and Scopus databases. Studies will be assessed for risk of bias using Cochrane’s RoB2 tool. Primary outcomes include a reduction in pain, measured quantitatively through pain scales. Secondary outcomes encompass quality of life, hospital length of stay, and opioid equivalents used.

**Discussion:** Existing literature underscores the therapeutic potential of amino acids, yet there still lacks a systematic review comparing the overall effects of different amino acid treatments for vaso-occlusive crises in patients with SCD. This review aims to serve as a valuable resource for clinicians, offering insights into amino acid interventions as alternatives or supplements to opioid treatments. Additionally, it seeks to encourage further randomized clinical trials, contributing to an informed clinical use of amino acids for pain management in SCD. Ultimately, the findings aim to enhance the understanding of the therapeutic effects of essential amino acids on pediatric patients with SCD, facilitating evidence-based clinical decisions.

## 3 Introduction

Sickle Cell Disease (SCD) affects over 100,000 individuals in the United States and 20 million globally, manifesting through acute and chronic pain syndromes.^1^ It is more prevalent among people of African, Mediterranean, Middle Eastern, and South Asian descent. The disease is caused by mutations in the beta hemoglobin genes, leading to the production of sickle hemoglobin (HbS)^2^ This results in red blood cells that can become deformed in the setting of physical and oxidative stress. The sickle-shaped erythrocytes become stiff and can obstruct smaller blood vessels, reducing blood flow and oxygen delivery to tissues. This can cause multi-organ damage, chronic anemia, and severe pain crises.^3^ Continuous vaso-occlusion and inflammation can progressively damage organs, and infections can exacerbate the severity of SCD.^4^ Consequently, the median life expectancy for individuals with SCD in the United States is approximately 43 years.^5^

Furthermore, SCD patients experience disproportionately high rates of emergency department (ED) usage. On average, there are 222,612 SCD-related ED visits annually,^6^ translating to 220 visits per 100 SCD patients, compared to 42.7 visits per 100 people in the general population.^7^ Vaso-occlusive crises (VOC) are the most common complication seen in SCD, accounting for 79% to 91% of these emergency visits.^8^ The high frequency of emergency department visits and the severe complications associated with Sickle Cell Disease highlight the urgent need for improved management and treatment strategies.

Amino acids contribute to energy metabolism and the starvation response, serve as precursors for several biological molecules including neurotransmitters, and are crucial for cell signaling and homeostasis^9^. The potential of amino acids for clinical intervention has long been investigated for various diseases. Aspartate and asparagine, as well as the branched-chain amino acids, have been shown to have a role in cancer metabolism.^10,11^ Other amino acids including lysine, glutamic acid, tryptophan, and glycine may have therapeutic use in treating psychiatric diseases including anxiety, schizophrenia, and sleep.^12^ Of particular interest for the treatment of SCD are the amino acids glutamine, arginine, and citrulline.

Glutamine, the most abundant amino acid in the body, has been investigated extensively for its potential benefits for SCD. A Phase 3 clinical trial demonstrated that L-glutamine significantly reduced pain crises and hospitalizations in SCD patients.^13^ L-glutamine oral powder was approved for treatment in patients aged 5 and older in 2017.^14^ While the exact mechanism of L-glutamine in SCD is not known, studies have shown that L-glutamine increases NADH levels and reduces red blood cell adhesion to the endothelium,^15^ supporting its potential therapeutic effect.

Additionally, arginine has garnered attention as a potential treatment for SCD. Clinical trials have shown that arginine supplementation can increase NO levels, improve endothelial function,^16^ and reduce the frequency and severity of pain episodes in SCD patients, leading to a reduction in pain crisis frequency and hospitalizations.^17^ Other studies have confirmed that arginine supplementation enhances NO production and may improve overall vascular health in SCD patients.^18^

In addition to L-glutamine and arginine, citrulline is crucial for the urea cycle and boosts nitric oxide (NO) production by increasing the availability of L-arginine. NO increases cyclic guanosine monophosphate (cGMP) synthesis, which promotes vasodilation, reduces arterial resistance and improves blood flow.^19^ A 2001 pilot Phase II clinical trial showed that oral L-citrulline administration improved symptoms in SCD patients, increased plasma arginine levels, and normalized leukocyte and neutrophil counts,^20^ suggesting significant therapeutic benefits.

Despite significant advancements in medical research, FDA approved treatments for SCD remain limited. There are currently four FDA-approved pharmacological interventions for SCD – hydroxyurea (Droxia), L-glutamine (Endari), voxelotor (Oxbryta), and crizanlizuman (Adakveo), with the latter three being approved less than a decade ago.^21^ Hydroxyurea reduces the frequency of painful crises and the need for blood transfusions by increasing fetal hemoglobin levels and reducing the need for blood transfusions.^22^ L-glutamine helps to mitigate oxidative stress in red blood cells, thus decreasing the occurrence of sickle cell crises.^23^ Voxelotor works by increasing hemoglobin’s affinity for oxygen, preventing cell sickling,^24^ while crizanlizumab reduces vaso-occlusive crises by inhibiting cell adhesion within blood vessels.^25^ These treatments are critical tools in the management of SCD, yet the limited number of options underscores the need for continued research and development in this field.

Given the success of several amino acid therapies in addressing Sickle Cell Disease and the limited pharmacological interventions, it is imperative that amino acids be analyzed as a potential therapeutic avenue for SCD management. Understanding the efficacy of these interventions at a systematic level is crucial, which underscores the importance of this systematic review. By compiling and analyzing existing research, this review aims to provide a comprehensive evaluation of amino acid treatments, thereby guiding clinical practice and future research in SCD management.

## 4 Methods

In conducting this systematic review, we will follow the methodology outlined in the Cochrane Handbook for Systematic Reviews of Interventions. This systematic review protocol is reported in accordance with the Preferred Reporting Items for Systematic Reviews and Meta-Analysis (PRISMA-P) guide^26^ and has been registered in the International Prospective Register of Systematic Reviews (PROSPERO) [CRD42024557365]). Upon completion, this systematic review will be reported in alignment with the PRISMA 2020 statement

## 5 Eligibility Criteria

Studies that compare amino acid treatment to placebo or to standard care for sickle cell disease (SCD) patients experiencing pain crises will be included in the analysis. Studies with participants of all age ranges will be considered. We will consider studies conducted in either inpatient or outpatient settings. There are no limitations on the particular amino acid treatment employed, provided that it was used to prevent or treat pain crises associated with SCD. No restrictions will be applied to the nature of the exposure intervention, encompassing both observational studies, such as cohort studies, and controlled clinical trials. However, case reports will be excluded. We will consider studies of all dose ranges of amino acid treatment used in the analysis. The comparator group will be patients receiving placebo or standard pain management treatment such as analgesia. There will be no limitations on the study’s publication year and country of origin. We will only include studies that enrolled participants for the sole purpose of treating SCD pain crises. Studies written in other languages besides English will be excluded.

## 6 Search Strategy

To identify relevant studies, we will conduct a search of the PubMed, Embase, and Scopus databases. Our search strategies will utilize a combination of terms derived from the PICOS framework, as outlined in Supplemental Table 1.

## 7 Selection of Studies

The studies compiled via the search strategy will be screened for their inclusion in the review. Five authors (BZ, CB, SY, PP, and VD) will carry out the screening of the studies. The titles and abstracts will be the initial review with each study being reviewed by two independent authors. Studies that do not meet the eligibility criteria will be excluded. An independent author (OBM) will resolve any disagreements regarding study inclusion. Following this, each study in its entirety will be reviewed by two independent authors. Disagreements will be handled by an independent author (OBM). A PRISMA flow chart will be included to overview the selection process.

## Data Availability

No datasets were generated or analysed during the current study. All relevant data from this study will be made available upon study completion.

## 8 Data Extraction and Management

Two review authors will independently extract data using Covidence prior to entering information into Review Manager4 (RevMan5.4). A third author will adjudicate in case of disagreement. We will extract the following information:

- Bibliographic data: title, authors, years, date of publication
- Methods: aims of study, study design, method of recruitment
- Characteristics of participants: age, gender, duration of pain, quality of life
- Treatment used: description of amino acid treatment
- Description of control group intervention: opioid therapy vs. amino acid treatment
- Outcomes of Interest: pain, quality of life, hospital length of stay, opioid equivalents used

## 9 Outcome Measures

### Primary Outcomes

Reduction in pain upon administration of amino acid treatment during pain crises will be the primary outcome. Progression in pain levels will be quantitatively recorded through the assessment of pain measures on the Visual Analogue Scale (VAS), Faces Pain Scale (FPS), and Numerical Rating Scale (NRS).

### 10 Secondary Outcomes

The quality of life will be assessed as a secondary outcome. Progression in quality of life will be quantified through measures of the Quality-of-Life Scale (QOLS0, 36-Item Short Form survey (SF-36), and other quantitative measures of quality of life. The length of hospital stay and the amount of opioid equivalents used will also be compared and analyzed as secondary outcomes.

## 11 Assessment of Risk

In accordance with Cochrane’s

Risk-of-Bias assessment tool (RoB2),^27^ authors will assess the following bias domains:

- Bias from randomization due to allocation sequence and treatment baseline differences
- Bias from subject and researcher blinding and potential treatment deviations due to this bias
- Bias from incomplete outcome data and potential non-randomization as a result
- Bias from differential measurement of studies’ outcomes
- Bias from researchers’ selectivity of reported results

All studies that meet the inclusion criteria requirement to be selected will be assessed by two authors independently in each risk category. Each author will evaluate the study in the risk category as: low risk of bias, high risk of bias, and unknown. In case of disagreement, a third author will mediate a discussion to reach a conclusion.

## 12 Data Analysis and Synthesis

If the trials in the included studies contain data that is similar in regards to participants, settings, intervention, comparison, and outcome measure, a meta-analysis will be conducted. In this case, the RevMan5.4 software will be used. Assuming studies will have different outcome measurements, the standardized mean difference with a corresponding confidence interval of 95% will be presented. This meta-analysis will then be illustrated with a forest plot. To determine the variation across the study due to heterogeneity, rather than chance, we will use the I^2^ statistic, where 0-40%, 30-60%, 50-90%, and 75-100% will correlate respectively to insignificant, moderate, substantial, and considerable heterogeneity. If data is not sufficiently similar, or the review contains other significant findings, subgroup analysis will be performed. To evaluate the risk of bias or publication bias from excluding studies, sensitivity analysis will be performed. The Egger test and funnel plots will be used. A narrative summary will accompany the tabulated and/or charted results and will describe how the results relate to the review’s objective of analyzing the different therapeutic effects of essential amino acids on patients.

## 13 Assessing the Certainty of Evidence

In assessing the certainty of evidence, we will adhere to the Grading of Recommendations, Assessment, Development, and Evaluation (GRADE) approach. We will utilize the following criteria to classify into four levels of certainty:

- High: very confident that true effect is close to estimated effect
- Moderate: moderately confident in estimated effect
- Low: confidence in estimated effect is limited
- Very low: very little confidence in estimated effect We will utilize the GRADEpro software28 in accordance with the GRADE approach to create a “Summary of Findings” table to evaluate the evidence.

We will utilize the GRADEpro software^28^ in accordance with the GRADE approach to create a “Summary of Findings” table to evaluate the evidence.

## 14 Discussions

This systematic review aims to identify the therapeutic effects of amino acid treatments in patients experiencing vaso-occlusive pain associated with Sickle Cell Disease through assessing pain, quality of life, hospital length of stay, and amount of opioid equivalent used post-administration of amino acid treatments.

Existing literature underscores the therapeutic potential of specific amino acids, such as glutamine, in treating vaso-occlusive pain crises in SCD patients. L-glutamine has shown efficacy in reducing the frequency of pain episodes and improving the overall health outcomes of SCD patients by reducing oxidative stress in red blood cells. However, despite promising results, there is a notable lack of comprehensive systematic reviews comparing the overall effects of amino acids on vaso-occlusive pain crises in this patient population.

This review aims to fill this gap by systematically compiling and analyzing existing research on the subject. By doing so, we seek to provide a valuable resource for clinicians, guiding them in prescribing amino acid pharmacological interventions as a supplement or alternative to opioid treatments for managing pain crises in individuals with SCD. Opioid treatments, while effective in pain management, are associated with significant risks and side effects. Therefore, alternative treatments such as amino acids could offer a safer, adjunctive approach to pain management in SCD.

Moreover, this review anticipates promoting further randomized clinical trials to systematically study the effects of amino acids on vaso-occlusive pain crises. Highlighting the potential benefits and gaps in the current research could pave the way for expanded and informed clinical use of amino acid interventions. This is particularly important given the limited FDA-approved treatments for SCD, and the addition of amino acid treatments could potentially enhance the therapeutic methods available for managing SCD pain crises.

In conclusion, this novel systematic review will serve as an important tool for future references in addressing pain with SCD, potentially leading to improved patient outcomes and quality of life. It aims to provide a comprehensive understanding of the role of amino acids in SCD management, thereby influencing clinical practices and future research directions in this field.

## 16 Supplemental Data

**S1 Table.**
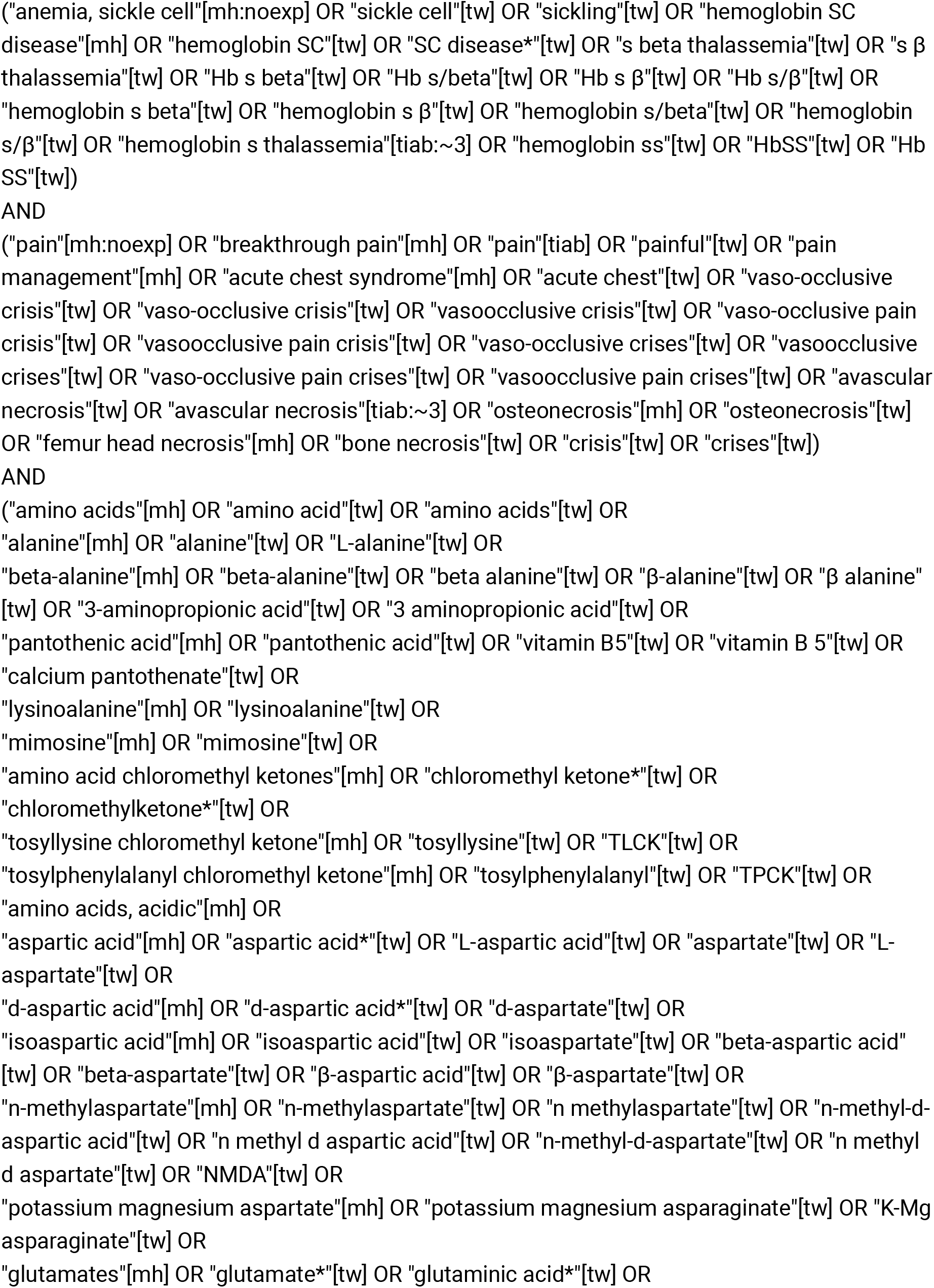
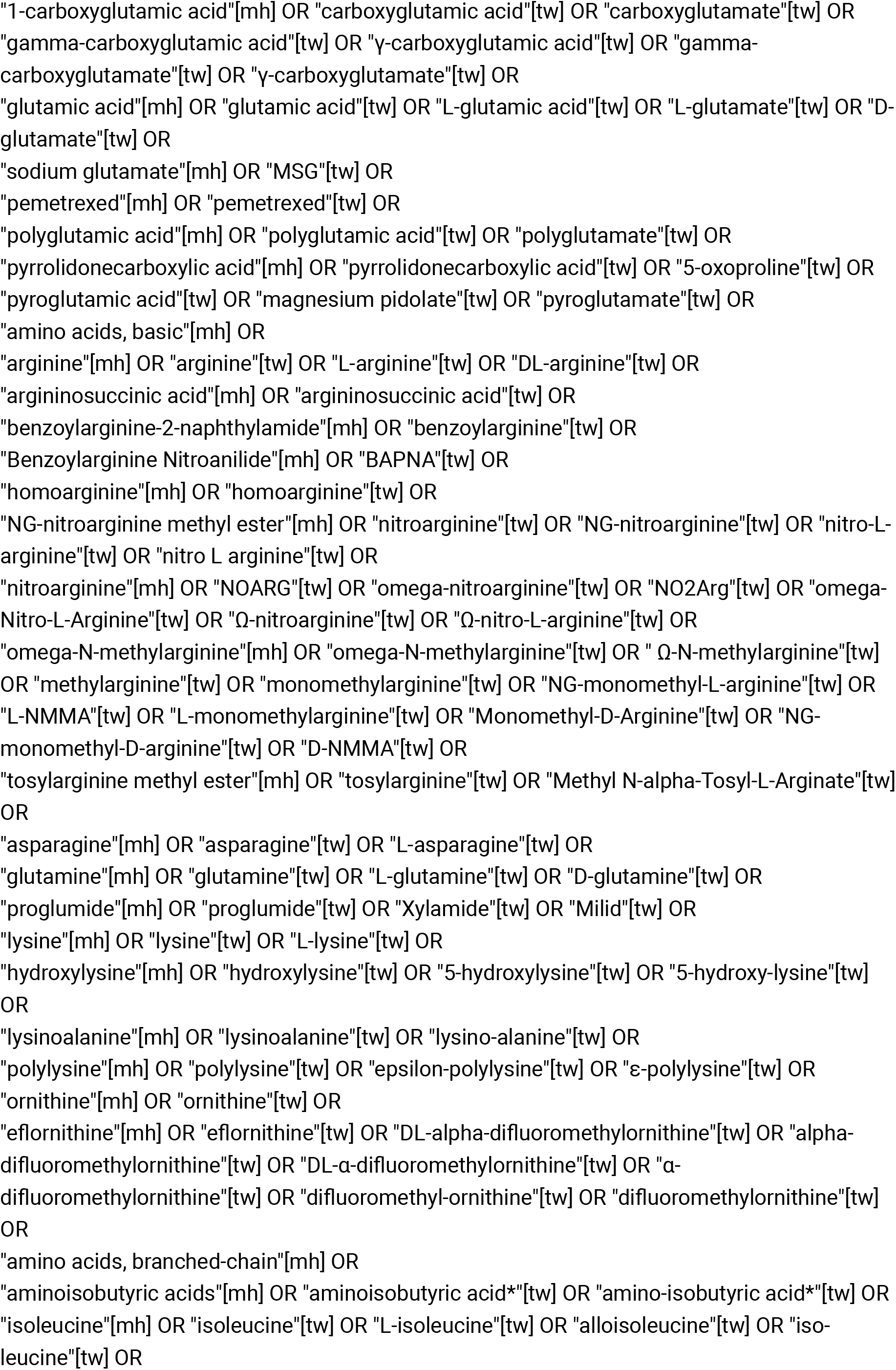
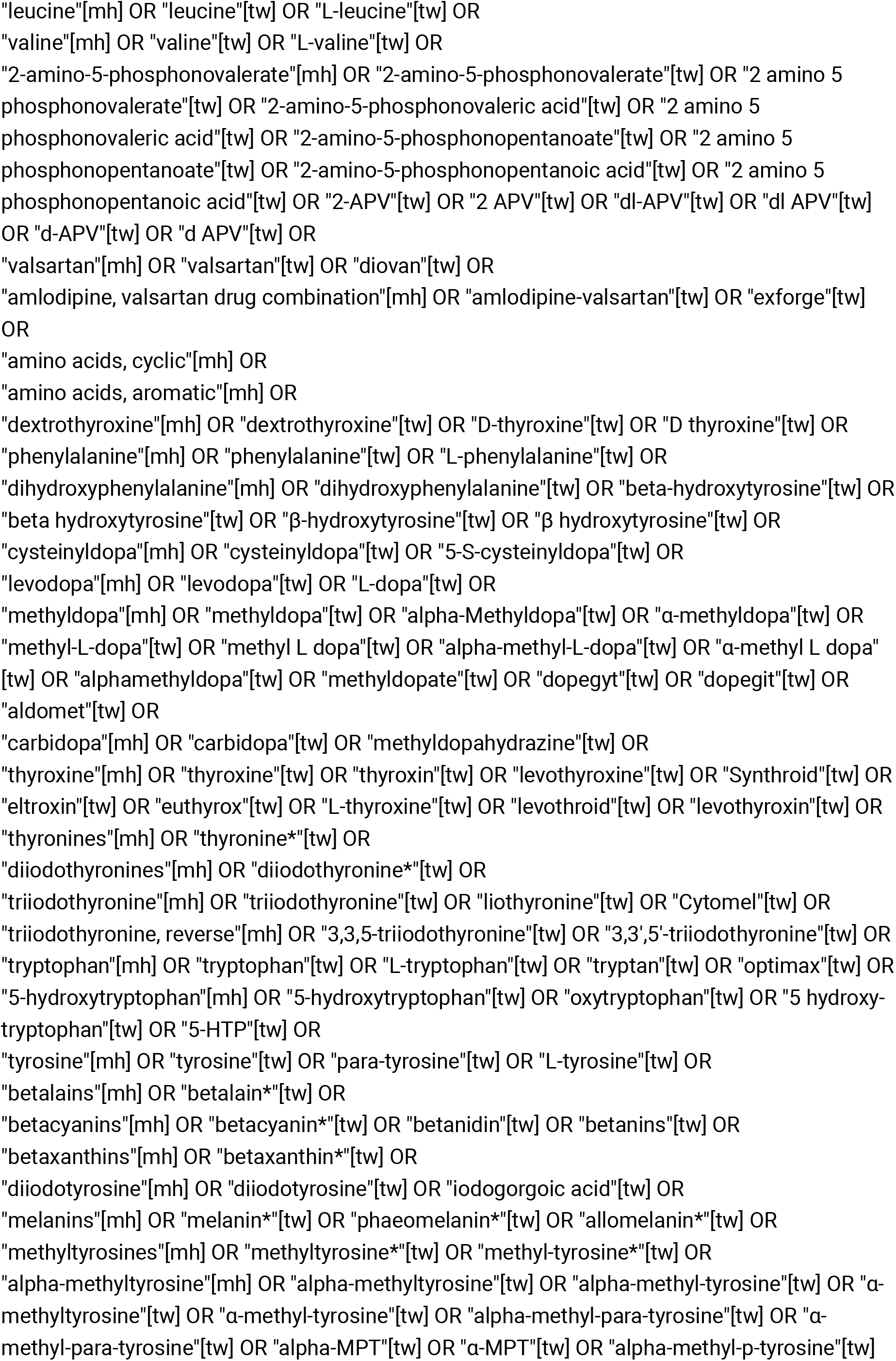
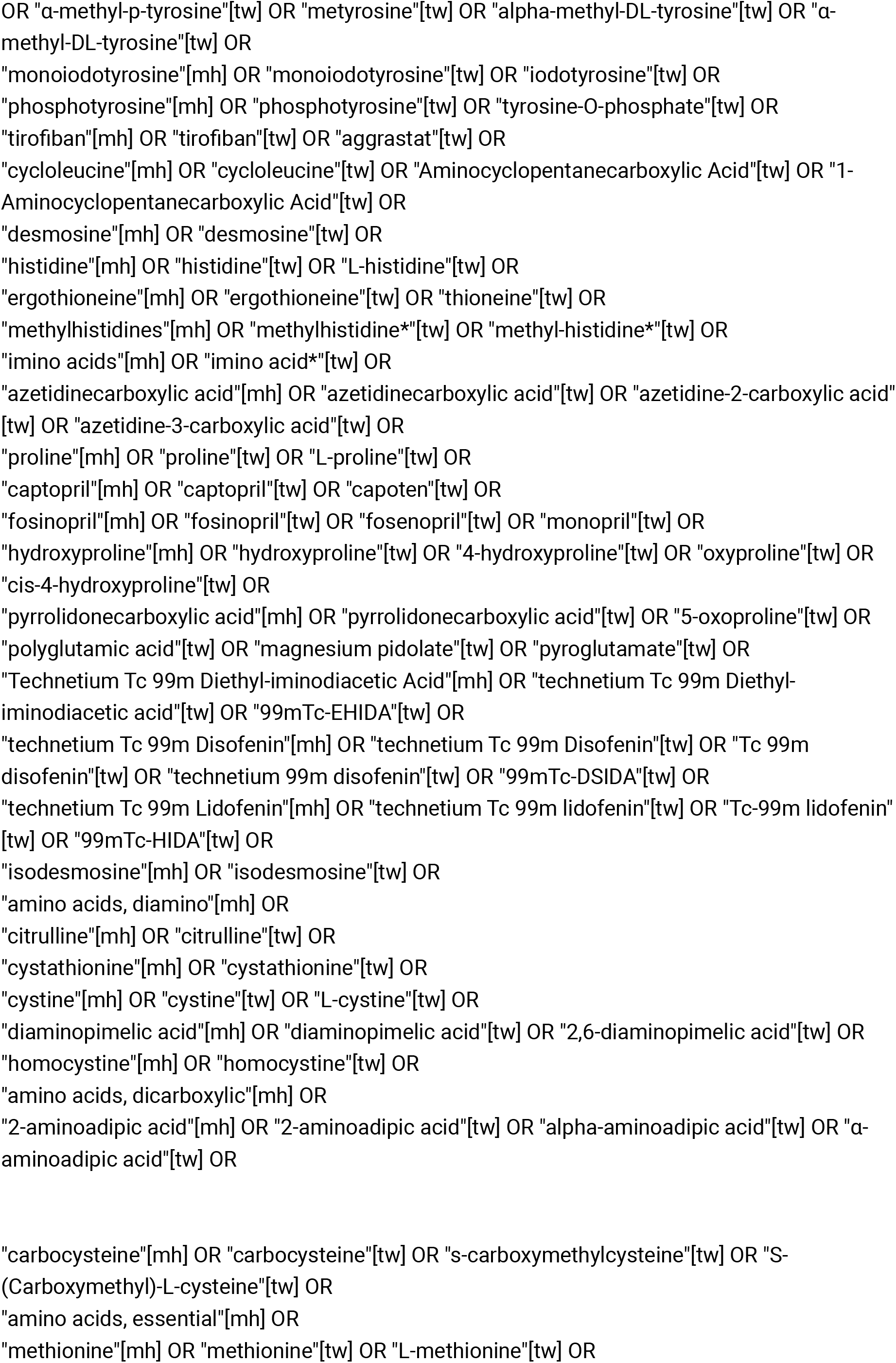
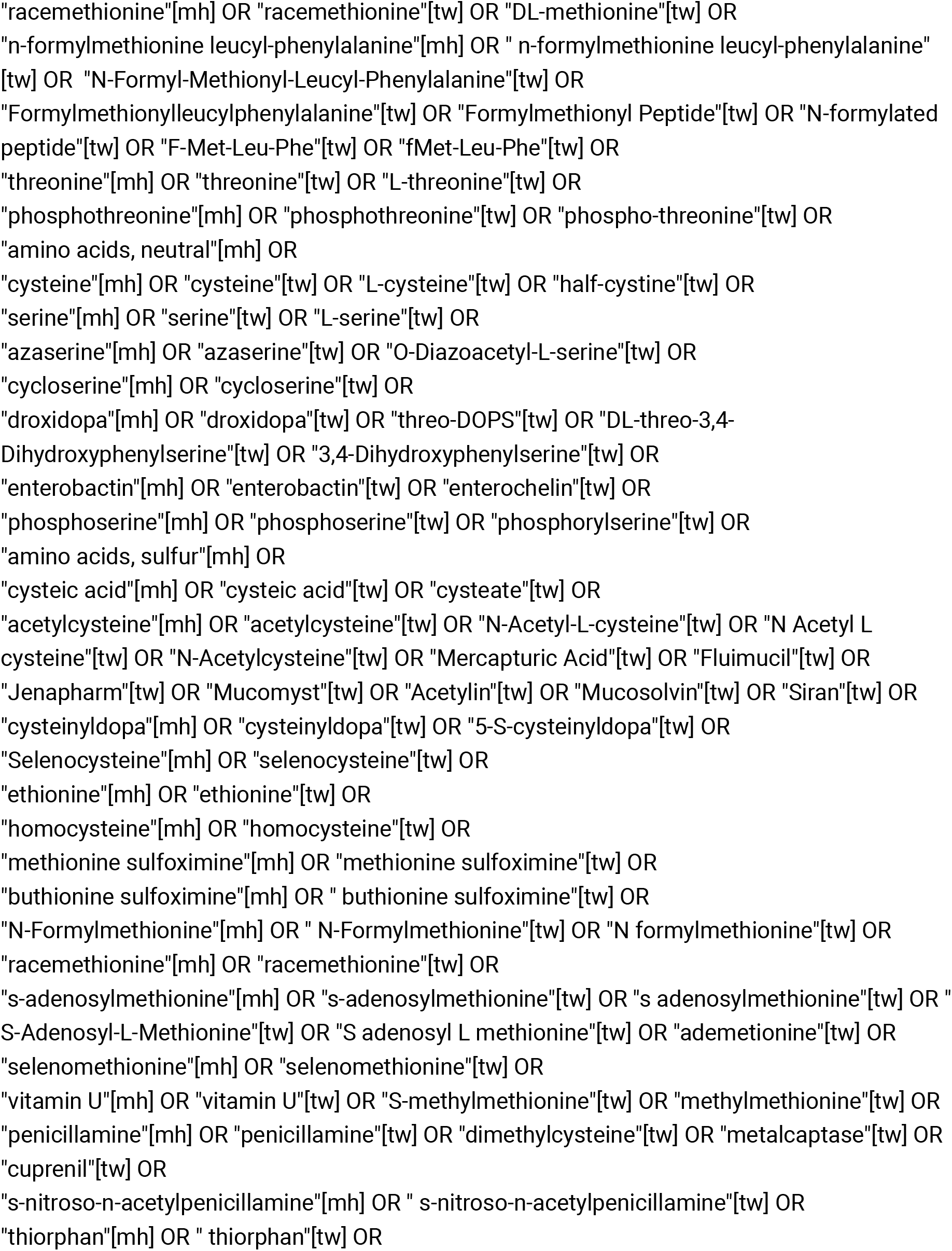

